# Sex differences in the associations of human milk oligosaccharides with height and weight in breastfed Ugandan children

**DOI:** 10.1101/2024.03.28.24305032

**Authors:** Tonny Jimmy Owalla, Victor Irungu Mwangi, Sara Moukarzel, Emmanuel Okurut, Chloe Yonemitsu, Lars Bode, Thomas G. Egwang

## Abstract

**Background & Objective:** Human milk oligosaccharides (HMOs) have been associated with several child growth metrics, but there was no difference in the associations according to child sex. Here, we present clinic-based cross-sectional data on the relationship between specific HMOs in the milk of Ugandan mothers and growth in their breastfed children as a group and as males and females separately.

**Method:** Human milk samples were manually collected from 127 lactating mothers. Levels of human milk oligosaccharides (HMOs) were analyzed by high-performance liquid chromatography (HPLC). Children’s weight and length were measured using a portable digital scale and length board, respectively. Weight-for-age (WAZ), height-for-age (HAZ) and weight for height (WHZ) Z scores were calculated. The relationships between HMOs and untransformed weights and heights and between WAZ, HAZ and WHZ subcategories were analyzed for all children and for males and females separately by Spearman’s correlation and Kruskal‒Wallis and Mann‒Whitney U tests.

**Results:** There were positive and negative correlations between the concentrations of specific HMOs and the heights and weights of children according to mothers’ secretor status. Tall infant stature was associated with higher concentrations of 6’SL, LSTc, DFLNH, DSLNH, LNnT and total HMOs in secretor or nonsecretor mothers; short infant stature was associated with higher concentrations of 3FL and DFLac; overweight was associated with higher concentrations of 6’SL, LSTc and LNnT in secretor and nonsecretor mothers; and normal weight was associated with higher concentrations of 3’FL. The associations between maternal HMO levels and childrens heights and weights or between the LAZ, WAZ and WHZ subcategories significantly differed between male and female children.

**Conclusions:** There are sex differences in the associations between high concentrations of some HMOs and stature and weight in Ugandan children born to secretor and nonsecretor mothers.

**Highlights:**

- This study showed that some HMOs are associated with child growth metrics in a sex-specific pattern.
- Four HMOs (3FL, DSLNT, DFLac, and 3’SL) were positively correlated with the height/length of the children.
- There were significant positive correlations between height/length and 3FL and DSLNT in only female children and between height/length and DFLac in only male children.
- DSLNT, DFLac and LSTb were positively correlated with weight in female and male children respectively.

**Plain Language Summary:** Human milk oligosaccharides (HMOs) have been associated with growth parameters of children in the Americas, Europe and Asia. Only two such studies have been conducted in Africa. None of the studies investigated the relationship between child sex and HMO composition and growth interactions. We examined the relationship between 19 well-characterized HMOs and growth metrics and also dissected the data by infant sex. Our results show a positive correlation between some HMOs and growth indices in either female or male children only. This highlights the impact of sex differences in the relationship between specific HMOs and growth measures in children. This evidence, if validated, could inform future nutritional interventions involving combinations of HMOs as food supplements that are equally effective for both male and female children.

## Background

Human milk contains an assortment of nutrients and bioactive molecules [1] and constitutes the best source of nutrition for infants, especially during the first 6 months of life [2,3]. Human milk components provide nourishment and play important developmental and protective functions. Compared with breastfed infants, nonbreastfed infants and children have a 14-fold greater risk of all-cause mortality and 8.6-fold higher infection-related mortality [2]. The benefits of breastfeeding can be partly attributed to human milk oligosaccharides (HMOs), a family of more than 150 nondigestible unconjugated carbohydrate molecules and the third most abundant biomolecule in human milk [4]. HMOs contain lactose at the reducing end that can be elongated by one or more disaccharides, such as N-acetyl-glucosamine and galactose [4,5]. Either lactose alone or elongated HMOs can be fucosylated at alpha-1-2/3/4-linkages and/or sialylated at alpha-2-3/6-linkages to form a variety of structurally distinct HMOs [4,5].

There is strong evidence that HMOs play a critical role in protecting children against infection through modulation of the gut microbiota and the immune system [6] and by serving as decoy receptors that block the epithelial attachment of various classes of pathogens [4,7,8]. These HMO effects are composition- and structure-specific [6,9]. Population-based prospective studies of mother–infant pairs showed that maternal milk containing high levels of specific α1-2-linked fucosyloligosaccharides was associated with a reduced risk of diarrhea from Campylobacter, cholera, and stable toxins of enteropathogenic *Escherichia coli* in breastfed children [10]. Infections caused by viral diarrhea pathogens such as norovirus and rotavirus diarrhea occurred less often in children whose mothers’ milk contained high levels of specific fucose-bound and sialic acid-bound oligosaccharides [11]. Remarkably, Lactodifucotetraose (LDFT) inhibited the release of proinflammatory proteins RANTES and sCD40L and platelet-induced inflammation [12], while 2’FL reduced mucosal-associated inflammation [13] in breastfed children. A clinical trial of 2’FL supplementation in infant formula confirmed its anti-inflammatory activity [14]. In addition to known anti-infective and anti-inflammatory functions, HMOs, in general, and sialic acid-bound HMOs, such as 3’SL and 6’SL, in particular, play significant roles in cognitive and brain development [15,16,17,18,19].

Studies in various countries have demonstrated an association between HMO concentrations or uptake and growth in children [20,21,22,23,24,25]. Lagstrom et al. (2020) showed that the concentrations of lacto-N-neo-tetraose (LNnT) and 2’FL were inversely and directly associated with height and weight, respectively, among children of Finnish secretor mothers [20]. In USA mother–infant dyads, lacto-N-fucopentaose I (LNFPI) was associated with lower infant weight, whereas disialyl-lacto-N-tetraose (DSLNT) and LNFPII were associated with greater fat mass [21]. Infant intakes of 3-fucosyllactose, 3’-sialyllactose (3’SL), 6’-sialyllactose (6’SL), DSLNT, disialyllacto-N-hexaose (DSLNH), and total sialylated HMOs were positively associated with infant growth over the first 6 months of life [22]. A Gambian study involving 33 mother–infant pairs concluded that 3’SL was a good indicator of infant weight-for-age [23], and several individual HMOs were associated with growth and development in a large study involving more than 600 Malawian mother–infant pairs [24]. Commercially available HMOs in infant formula provided growth benefits comparable to those of breastfeeding in studies carried out in non-African countries [26,27,28].

There is a paucity of studies conducted in Africa that have investigated the relationship between HMOs and growth in breastfeeding children, and the outcomes of these studies are contradictory [23,24]. It was of interest to investigate the relationship between specific HMOs in the milk of Ugandan mothers and growth outcomes in their breastfed children. Here, we report on sex-dependent associations between various HMOs and growth in breastfed Ugandan children.

## Methods

### Study design and population

The study population was a subset of a cohort of 400 mother–child pairs under 5 years old who were participating in a prospective longitudinal study investigating the effectiveness of a malaria vector control intervention in Abwokodia Parish, Katakwi district in northeastern Uganda. Details of the study site have been reported elsewhere [29]. This breast milk substudy involved a clinic-based cross-sectional survey of breastfeeding mother–infant pairs. All breastfeeding infants less than two years old and their respective breastfeeding mothers were considered eligible for the study upon consent by one or both parents, as applicable. One hundred and twenty-seven (127) breastfeeding mother–infant pairs were identified and included in the study.

### Participant recruitment and anthropometric measurements

The study was conducted in March 2018. Mothers of all breastfeeding children who were participating in an ongoing longitudinal study were mobilized by a social scientist with the help of village health teams. Mother–child pairs visited the study clinic at St. Anne Health Center III for screening and enrollment. The objectives of the study were explained to the mothers, and informed consent was obtained. Milk samples were collected from consenting mothers. Child weight and length and other anthropometric data for both mothers and children were measured using a portable digital scale (Seca 334) and length board (Seca 417), respectively.

### Human milk sampling and treatment

A single 5-mL sample of human milk was collected from each lactating mother by manual expression with the help of a senior midwife who is an experienced lactation counsellor. Milk samples were collected into sterile 50-mL Falcon tubes and immediately frozen in dry ice. Milk samples were subsequently transported on dry ice to Med Biotech Laboratories in Kampala, where they were stored at -20°C before being air-freighted on dry ice to the University of California San Diego for HMO analysis.

### HMO Extraction, Analysis, and Secretor Status Determination

Analysis of 19 well-characterized and most abundant HMOs was performed at the University of California, San Diego, as previously described [30]. Briefly, human milk samples were analyzed by HPLC after fluorescent derivatization. Raffinose was added to each milk sample as an internal standard for absolute quantification. The total concentration of HMOs was calculated as the sum of the specific oligosaccharides detected. The proportion of each HMO per total HMO concentration was calculated. The maternal phenotypic secretor status was determined by the relative abundance (secretor) or near absence (nonsecretor) of 2′FL and LNFP-I in the respective milk samples.

### Statistical analyses

The weight and height of each infant was used to calculate weight-for-age (WAZ), weight-for-length (WFL) and height-for-age (HAZ) Z scores based on the standards of the World Health Organization [31]. These scores were also grouped into categorical variables. Height was subcategorized as short, normal or tall, while weight was subcategorized as underweight, normal or overweight. Statistical analysis was performed using GraphPad Prism version 9.0.2 and STATA version 15. The Shapiro‒Wilk test was used to evaluate the distribution of variables, including oligosaccharide concentrations. Nonparametric comparisons were made using the Mann‒Whitney U test for paired samples or the Kruskal‒Wallis test for groups. Exploratory analyses were performed to compare median oligosaccharide concentrations according to children’s LAZ, WAZ and WHZ subcategories. The results are presented as box-plot graphs. Correlations between the oligosaccharide concentrations and the children’s weight or height were assessed using the Spearman rank test with 95% confidence intervals. Significance was defined as a *P* value less than .05.

## Results

### Characteristics of the study participants

The study population comprised 127 breastfeeding mother–child pairs from Katakwi District in northeastern Uganda. The mothers had a mean age of 26.6 years (range 15-46), with a postpartum mean BMI of 23.3 kg/m^2^ (18.7-31.8). Most of the mothers were multiparous (> 78%) and had breastfed for 3-103 weeks. The mean age, weight, and height/length of the children were 46 weeks (range 3-110), 8.8 kg (range 5-16), and 70.5 cm (range 50-98), respectively, and 52% were females.

### Correlation between HMO concentrations and heights of all children

We undertook Spearman’s correlation analysis to assess the effect of specific HMOs in mothers’ milk on the height or length of breastfed children. These results are presented in **Table 1**. There was a significant positive correlation between the height/length of children and four HMOs, namely, 3FL for all mothers combined and secretor mothers but not nonsecretor mothers; DSLNT for nonsecretor mothers only; DFLac for secretor mothers and nonsecretor mothers; and 3’SL for all mothers combined, secretor mothers and nonsecretor mothers. There was a significant negative correlation between heigh/length in children for 8 HMOs, namely, 6’SL, LSTc and DSLNH for all mothers combined; secretor mothers and non-secretor mothers; LNnT, LNH, FLNH for all mothers combined and secretor mothers; LNFP I for all mothers combined; and FDSLNH for nonsecretor mothers only. However, there was no significant correlation between the height or length of infants and children of all mothers and the diversity of 7 out of 19 HMOs, total HMOs, HMO-bound sialic acid, HMO-bound fucose, regardless of the secretor status.

**TABLE 1.**
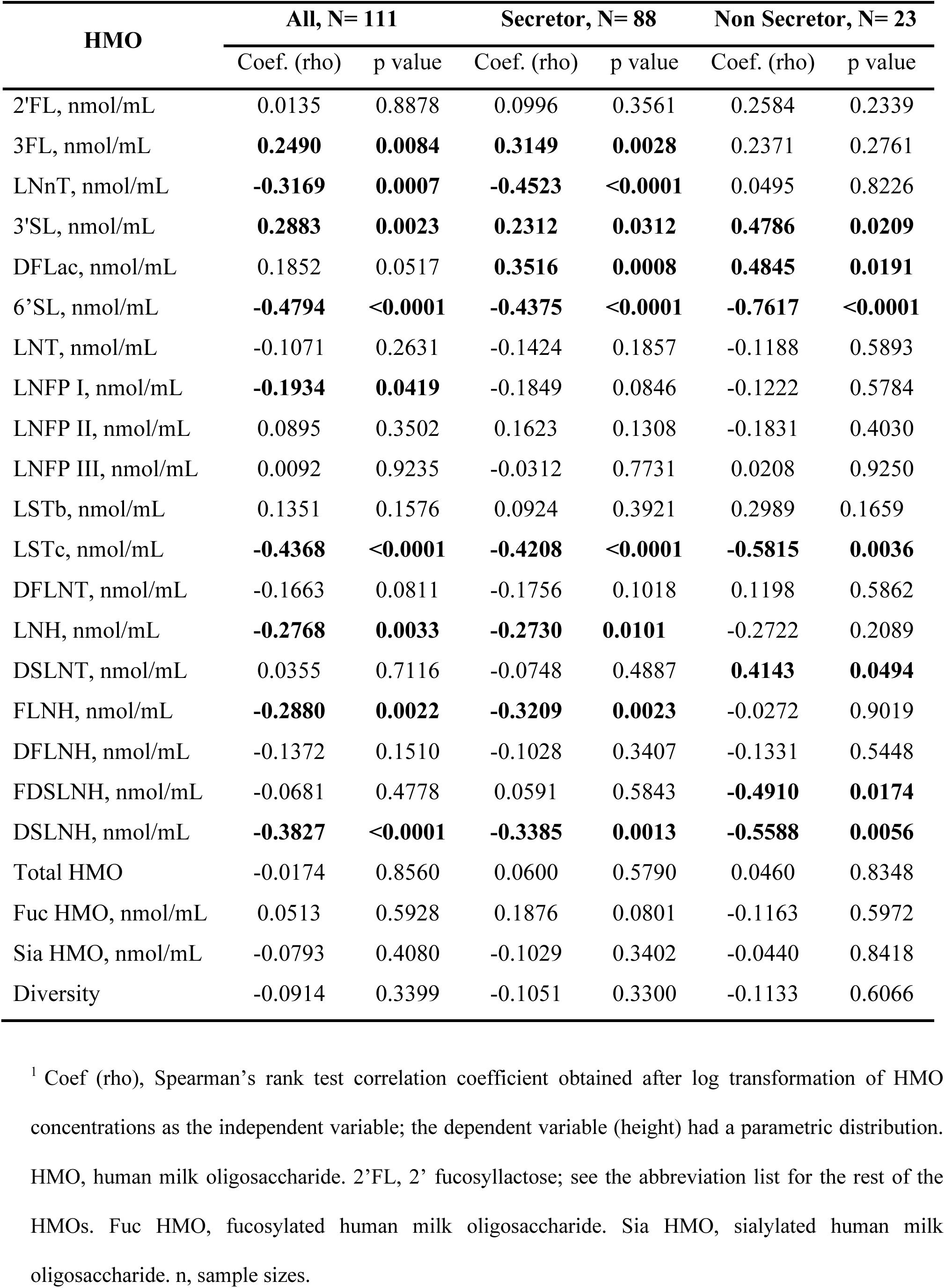
Correlations between HMOs and the heights of all children^1^.

| HMO | All, N= 111 |  | Secretor, N= 88 |  | Non Secretor, N= 23 |  |
| --- | --- | --- | --- | --- | --- | --- |
|  | Coef. (rho) | p value | Coef. (rho) | p value | Coef. (rho) | p value |
| 2'FL, nmol/mL | 0.0135 | 0.8878 | 0.0996 | 0.3561 | 0.2584 | 0.2339 |
| 3FL, nmol/mL | **0.2490** | **0.0084** | **0.3149** | **0.0028** | 0.2371 | 0.2761 |
| LNnT, nmol/mL | **-0.3169** | **0.0007** | **-0.4523** | **<0.0001** | 0.0495 | 0.8226 |
| 3'SL, nmol/mL | **0.2883** | **0.0023** | **0.2312** | **0.0312** | **0.4786** | **0.0209** |
| DFLac, nmol/mL | 0.1852 | 0.0517 | **0.3516** | **0.0008** | **0.4845** | **0.0191** |
| 6'SL, nmol/mL | **-0.4794** | **<0.0001** | **-0.4375** | **<0.0001** | **-0.7617** | **<0.0001** |
| LNT, nmol/mL | -0.1071 | 0.2631 | -0.1424 | 0.1857 | -0.1188 | 0.5893 |
| LNFP I, nmol/mL | **-0.1934** | **0.0419** | -0.1849 | 0.0846 | -0.1222 | 0.5784 |
| LNFP II, nmol/mL | 0.0895 | 0.3502 | 0.1623 | 0.1308 | -0.1831 | 0.4030 |
| LNFP III, nmol/mL | 0.0092 | 0.9235 | -0.0312 | 0.7731 | 0.0208 | 0.9250 |
| LSTb, nmol/mL | 0.1351 | 0.1576 | 0.0924 | 0.3921 | 0.2989 | 0.1659 |
| LSTc, nmol/mL | **-0.4368** | **<0.0001** | **-0.4208** | **<0.0001** | **-0.5815** | **0.0036** |
| DFLNT, nmol/mL | -0.1663 | 0.0811 | -0.1756 | 0.1018 | 0.1198 | 0.5862 |
| LNH, nmol/mL | **-0.2768** | **0.0033** | **-0.2730** | **0.0101** | -0.2722 | 0.2089 |
| DSLNT, nmol/mL | 0.0355 | 0.7116 | -0.0748 | 0.4887 | **0.4143** | **0.0494** |
| FLNH, nmol/mL | **-0.2880** | **0.0022** | **-0.3209** | **0.0023** | -0.0272 | 0.9019 |
| DFLNH, nmol/mL | -0.1372 | 0.1510 | -0.1028 | 0.3407 | -0.1331 | 0.5448 |
| FDSLNH, nmol/mL | -0.0681 | 0.4778 | 0.0591 | 0.5843 | **-0.4910** | **0.0174** |
| DSLNH, nmol/mL | **-0.3827** | **<0.0001** | **-0.3385** | **0.0013** | **-0.5588** | **0.0056** |
| Total HMO | -0.0174 | 0.8560 | 0.0600 | 0.5790 | 0.0460 | 0.8348 |
| Fuc HMO, nmol/mL | 0.0513 | 0.5928 | 0.1876 | 0.0801 | -0.1163 | 0.5972 |
| Sia HMO, nmol/mL | -0.0793 | 0.4080 | -0.1029 | 0.3402 | -0.0440 | 0.8418 |
| Diversity | -0.0914 | 0.3399 | -0.1051 | 0.3300 | -0.1133 | 0.6066 |

<sup>1</sup> Coef (rho), Spearman's rank test correlation coefficient obtained after log transformation of HMO concentrations as the independent variable; the dependent variable (height) had a parametric distribution. HMO, human milk oligosaccharide. 2'FL, 2' fucosyllactose; see the abbreviation list for the rest of the HMOs. Fuc HMO, fucosylated human milk oligosaccharide. Sia HMO, sialylated human milk oligosaccharide. n, sample sizes.

### Correlations between HMO concentrations and height according to sex

There were differences between male and female children of secretor and nonsecretor mothers in terms of the correlation of specific HMOs with height (**Table 2**). First, there was a significant positive correlation between height and 3FL in only female children and DFLac in only male children of secretor but not nonsecretor mothers; and between height and DSLNT in only female children of nonsecretor mothers. Second, there were significant negative correlations between height and LNT, LNnT and HMO-bound fucose in female children and between FLNH and DSLNH in male children of secretor mothers. Third, there was a significant negative correlation with 6’SL in both male and female children of secretor mothers and nonsecretor mothers; with LSTc in both male and female children of secretor mothers but only in male children of nonsecretor mothers; and with DFLNH in male children of nonsecretor mothers. Finally, LNT and HMO-bound fucose did not significantly correlate with height in the children of all mothers combined, regardless of secretor status, when the children were not stratified by sex (**Table 1**). However, when children were stratified by sex, LNT and fucose-bound HMOs had a significant negative correlation with height for only female children of secretor mothers (**Table 2**).

**TABLE 2.**
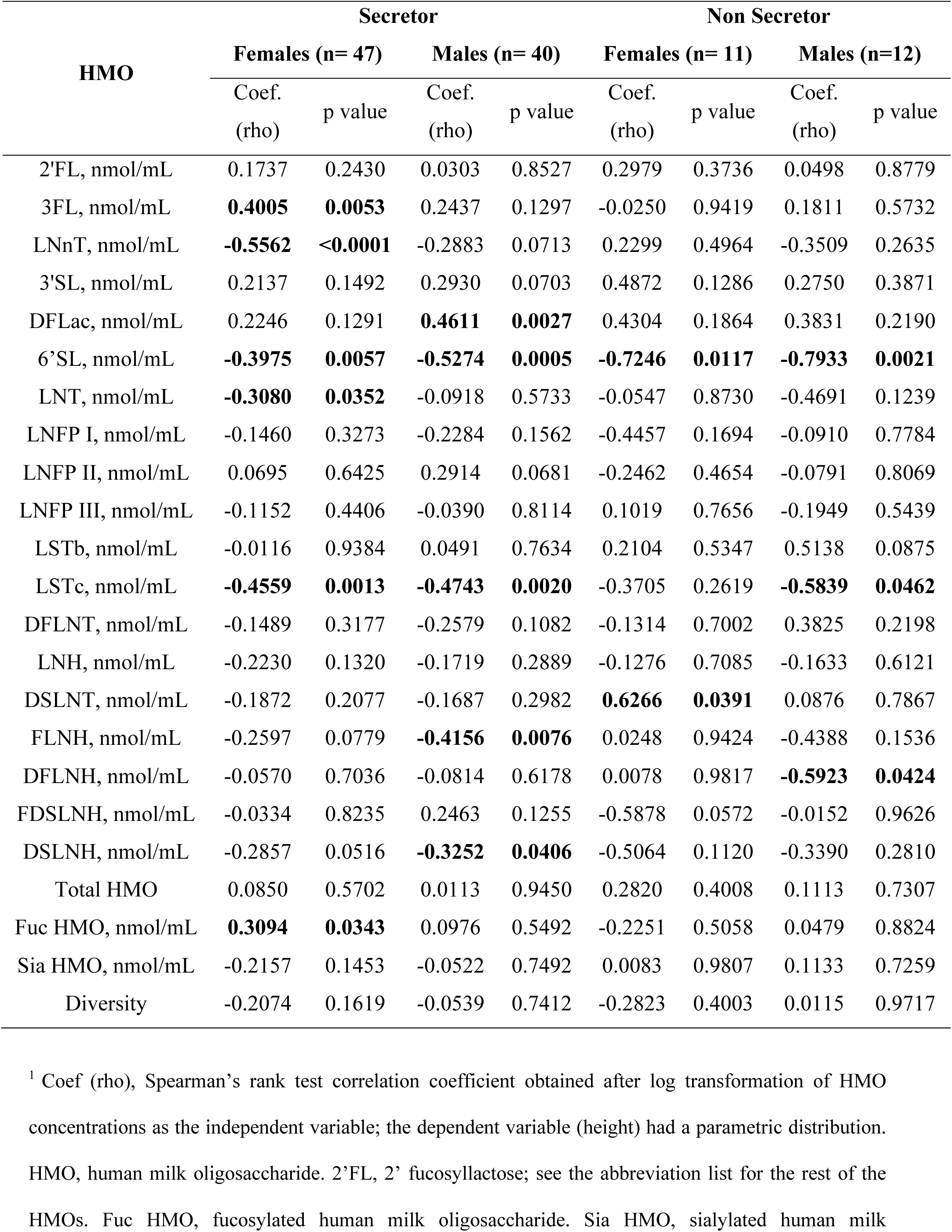
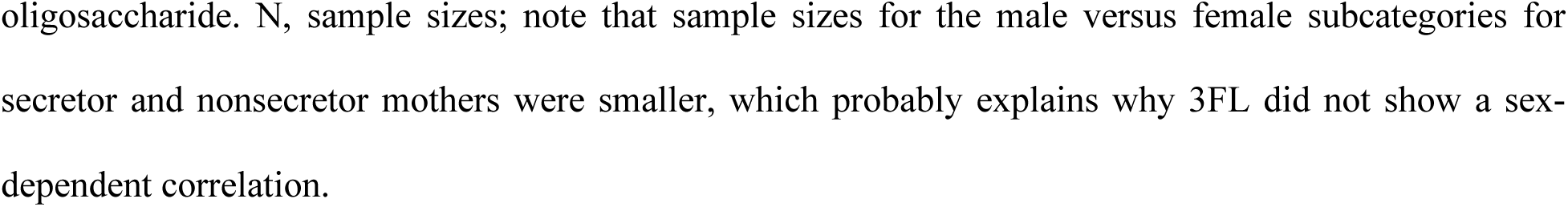
Correlations between HMOs and the height of female and male children^1^.

| HMO | Secretor |  |  |  | Non Secretor |  |  |  |
| --- | --- | --- | --- | --- | --- | --- | --- | --- |
|  | Females (n= 47) |  | Males (n= 40) |  | Females (n= 11) |  | Males (n=12) |  |
|  | Coef.<br>(rho) | p value | Coef.<br>(rho) | p value | Coef.<br>(rho) | p value | Coef.<br>(rho) | p value |
| 2'FL, nmol/mL | 0.1737 | 0.2430 | 0.0303 | 0.8527 | 0.2979 | 0.3736 | 0.0498 | 0.8779 |
| 3FL, nmol/mL | **0.4005** | **0.0053** | 0.2437 | 0.1297 | -0.0250 | 0.9419 | 0.1811 | 0.5732 |
| LNnT, nmol/mL | **-0.5562** | **<0.0001** | -0.2883 | 0.0713 | 0.2299 | 0.4964 | -0.3509 | 0.2635 |
| 3'SL, nmol/mL | 0.2137 | 0.1492 | 0.2930 | 0.0703 | 0.4872 | 0.1286 | 0.2750 | 0.3871 |
| DFLac, nmol/mL | 0.2246 | 0.1291 | **0.4611** | **0.0027** | 0.4304 | 0.1864 | 0.3831 | 0.2190 |
| 6'SL, nmol/mL | **-0.3975** | **0.0057** | **-0.5274** | **0.0005** | **-0.7246** | **0.0117** | **-0.7933** | **0.0021** |
| LNT, nmol/mL | **-0.3080** | **0.0352** | -0.0918 | 0.5733 | -0.0547 | 0.8730 | -0.4691 | 0.1239 |
| LNFP I, nmol/mL | -0.1460 | 0.3273 | -0.2284 | 0.1562 | -0.4457 | 0.1694 | -0.0910 | 0.7784 |
| LNFP II, nmol/mL | 0.0695 | 0.6425 | 0.2914 | 0.0681 | -0.2462 | 0.4654 | -0.0791 | 0.8069 |
| LNFP III, nmol/mL | -0.1152 | 0.4406 | -0.0390 | 0.8114 | 0.1019 | 0.7656 | -0.1949 | 0.5439 |
| LSTb, nmol/mL | -0.0116 | 0.9384 | 0.0491 | 0.7634 | 0.2104 | 0.5347 | 0.5138 | 0.0875 |
| LSTc, nmol/mL | **-0.4559** | **0.0013** | **-0.4743** | **0.0020** | -0.3705 | 0.2619 | **-0.5839** | **0.0462** |
| DFLNT, nmol/mL | -0.1489 | 0.3177 | -0.2579 | 0.1082 | -0.1314 | 0.7002 | 0.3825 | 0.2198 |
| LNH, nmol/mL | -0.2230 | 0.1320 | -0.1719 | 0.2889 | -0.1276 | 0.7085 | -0.1633 | 0.6121 |
| DSLNT, nmol/mL | -0.1872 | 0.2077 | -0.1687 | 0.2982 | **0.6266** | **0.0391** | 0.0876 | 0.7867 |
| FLNH, nmol/mL | -0.2597 | 0.0779 | **-0.4156** | **0.0076** | 0.0248 | 0.9424 | -0.4388 | 0.1536 |
| DFLNH, nmol/mL | -0.0570 | 0.7036 | -0.0814 | 0.6178 | 0.0078 | 0.9817 | **-0.5923** | **0.0424** |
| FDSLNH, nmol/mL | -0.0334 | 0.8235 | 0.2463 | 0.1255 | -0.5878 | 0.0572 | -0.0152 | 0.9626 |
| DSLNH, nmol/mL | -0.2857 | 0.0516 | **-0.3252** | **0.0406** | -0.5064 | 0.1120 | -0.3390 | 0.2810 |
| Total HMO | 0.0850 | 0.5702 | 0.0113 | 0.9450 | 0.2820 | 0.4008 | 0.1113 | 0.7307 |
| Fuc HMO, nmol/mL | **0.3094** | **0.0343** | 0.0976 | 0.5492 | -0.2251 | 0.5058 | 0.0479 | 0.8824 |
| Sia HMO, nmol/mL | -0.2157 | 0.1453 | -0.0522 | 0.7492 | 0.0083 | 0.9807 | 0.1133 | 0.7259 |
| Diversity | -0.2074 | 0.1619 | -0.0539 | 0.7412 | -0.2823 | 0.4003 | 0.0115 | 0.9717 |

<sup>1</sup> Coef (rho), Spearman's rank test correlation coefficient obtained after log transformation of HMO concentrations as the independent variable; the dependent variable (height) had a parametric distribution.
HMO, human milk oligosaccharide. 2'FL, 2' fucosyllactose; see the abbreviation list for the rest of the HMOs. Fuc HMO, fucosylated human milk oligosaccharide. Sia HMO, sialylated human milk
oligosaccharide. N, sample sizes; note that sample sizes for the male versus female subcategories for secretor and nonsecretor mothers were smaller, which probably explains why 3FL did not show a sex-dependent correlation.

### Correlations between HMO concentrations and weights of all children

We undertook Spearman’s correlation analysis to assess the association of specific HMOs in mothers’ milk with the weight of breastfed children (**Table 3**). There was a significant positive correlation between 3’SL and the weight of children of all mothers combined, nonsecretor mothers, and between DFLac and the weight of children of secretor mothers. In contrast, there were significant negative correlations between LNnT, LNH, and FLNH and the weights of children of all mothers combined and secretor mothers. Similarly, 6’SL, LSTc, and DSLNH correlated negatively with the weights of children of all mothers combined, with secretor and nonsecretor mothers. However, there were no correlations between total HMOs, HMO-bound sialic acid, or HMO-bound fucose, and weight of the children of mothers, regardless of their secretor status.

**TABLE 3.**
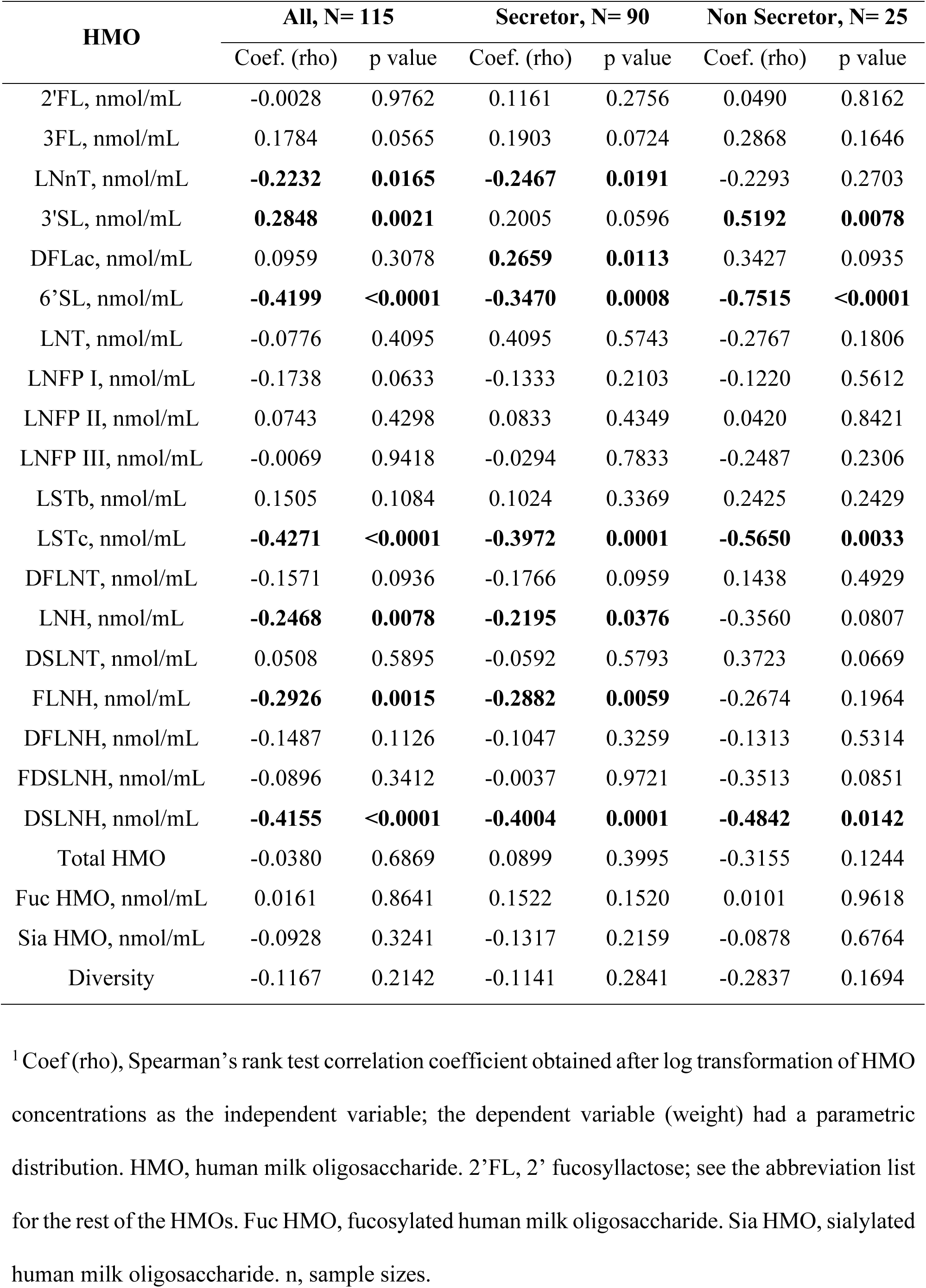
Correlations between HMOs and the weights of all children^1^.

| HMO | All, N= 115 |  | Secretor, N= 90 |  | Non Secretor, N= 25 |  |
| --- | --- | --- | --- | --- | --- | --- |
|  | Coef. (rho) | p value | Coef. (rho) | p value | Coef. (rho) | p value |
| 2'FL, nmol/mL | -0.0028 | 0.9762 | 0.1161 | 0.2756 | 0.0490 | 0.8162 |
| 3FL, nmol/mL | 0.1784 | 0.0565 | 0.1903 | 0.0724 | 0.2868 | 0.1646 |
| LNnT, nmol/mL | **-0.2232** | **0.0165** | **-0.2467** | **0.0191** | -0.2293 | 0.2703 |
| 3'SL, nmol/mL | **0.2848** | **0.0021** | 0.2005 | 0.0596 | **0.5192** | **0.0078** |
| DFLac, nmol/mL | 0.0959 | 0.3078 | **0.2659** | **0.0113** | 0.3427 | 0.0935 |
| 6'SL, nmol/mL | **-0.4199** | **<0.0001** | **-0.3470** | **0.0008** | **-0.7515** | **<0.0001** |
| LNT, nmol/mL | -0.0776 | 0.4095 | 0.4095 | 0.5743 | -0.2767 | 0.1806 |
| LNFP I, nmol/mL | -0.1738 | 0.0633 | -0.1333 | 0.2103 | -0.1220 | 0.5612 |
| LNFP II, nmol/mL | 0.0743 | 0.4298 | 0.0833 | 0.4349 | 0.0420 | 0.8421 |
| LNFP III, nmol/mL | -0.0069 | 0.9418 | -0.0294 | 0.7833 | -0.2487 | 0.2306 |
| LSTb, nmol/mL | 0.1505 | 0.1084 | 0.1024 | 0.3369 | 0.2425 | 0.2429 |
| LSTc, nmol/mL | **-0.4271** | **<0.0001** | **-0.3972** | **0.0001** | **-0.5650** | **0.0033** |
| DFLNT, nmol/mL | -0.1571 | 0.0936 | -0.1766 | 0.0959 | 0.1438 | 0.4929 |
| LNH, nmol/mL | **-0.2468** | **0.0078** | **-0.2195** | **0.0376** | -0.3560 | 0.0807 |
| DSLNT, nmol/mL | 0.0508 | 0.5895 | -0.0592 | 0.5793 | 0.3723 | 0.0669 |
| FLNH, nmol/mL | **-0.2926** | **0.0015** | **-0.2882** | **0.0059** | -0.2674 | 0.1964 |
| DFLNH, nmol/mL | -0.1487 | 0.1126 | -0.1047 | 0.3259 | -0.1313 | 0.5314 |
| FDSLNH, nmol/mL | -0.0896 | 0.3412 | -0.0037 | 0.9721 | -0.3513 | 0.0851 |
| DSLNH, nmol/mL | **-0.4155** | **<0.0001** | **-0.4004** | **0.0001** | **-0.4842** | **0.0142** |
| Total HMO | -0.0380 | 0.6869 | 0.0899 | 0.3995 | -0.3155 | 0.1244 |
| Fuc HMO, nmol/mL | 0.0161 | 0.8641 | 0.1522 | 0.1520 | 0.0101 | 0.9618 |
| Sia HMO, nmol/mL | -0.0928 | 0.3241 | -0.1317 | 0.2159 | -0.0878 | 0.6764 |
| Diversity | -0.1167 | 0.2142 | -0.1141 | 0.2841 | -0.2837 | 0.1694 |

<sup>1</sup> Coef (rho), Spearman's rank test correlation coefficient obtained after log transformation of HMO concentrations as the independent variable; the dependent variable (weight) had a parametric distribution. HMO, human milk oligosaccharide. 2'FL, 2' fucosyllactose; see the abbreviation list for the rest of the HMOs. Fuc HMO, fucosylated human milk oligosaccharide. Sia HMO, sialylated human milk oligosaccharide. n, sample sizes.

### Correlations between HMO concentrations and weights of male and female children

There were differences between male and female children of secretor and nonsecretor mothers in terms of the correlation of specific HMOs with weight (**Table 4**). First, there was a significant positive correlation between weight in male children and DFLac levels in secretor mothers; between weight in male children and LSTb levels in nonsecretor mothers, and between weight in female children and DSLNT levels in nonsecretor mothers. In contrast, there was a significant negative correlation between weight in male children and FLNH levels in secretor mothers, between weight in male children and DFLNH and 2’FL levels in nonsecretor mothers, and between weight in female children and LNFPI levels in nonsecretor mothers. Third, there was a negative correlation between weight and 6’SL in male children of secretor mothers and in both male and female children of nonsecretor mothers; between weight and LSTc in both male and female children of secretor mothers and in only female children of nonsecretors; and between DSLNH in both male and female children of secretor mothers and in only male children of nonsecretor mothers. Finally, 5 HMOs had no significant correlation with weight in children of all mothers combined, regardless of secretor status, when children were not grouped by sex (**Table 3**). However, when the children were grouped by sex, these HMOs had significant sex-dependent correlations with weight in children of nonsecretor mothers; 2’FL and DFLNH had significant negative, and LSTb had a significant positive correlation correlations in male children; LNFP I and DSLNT had a significant negative and positive correlation in female children respectively (**Table 4**).

**TABLE 4.**
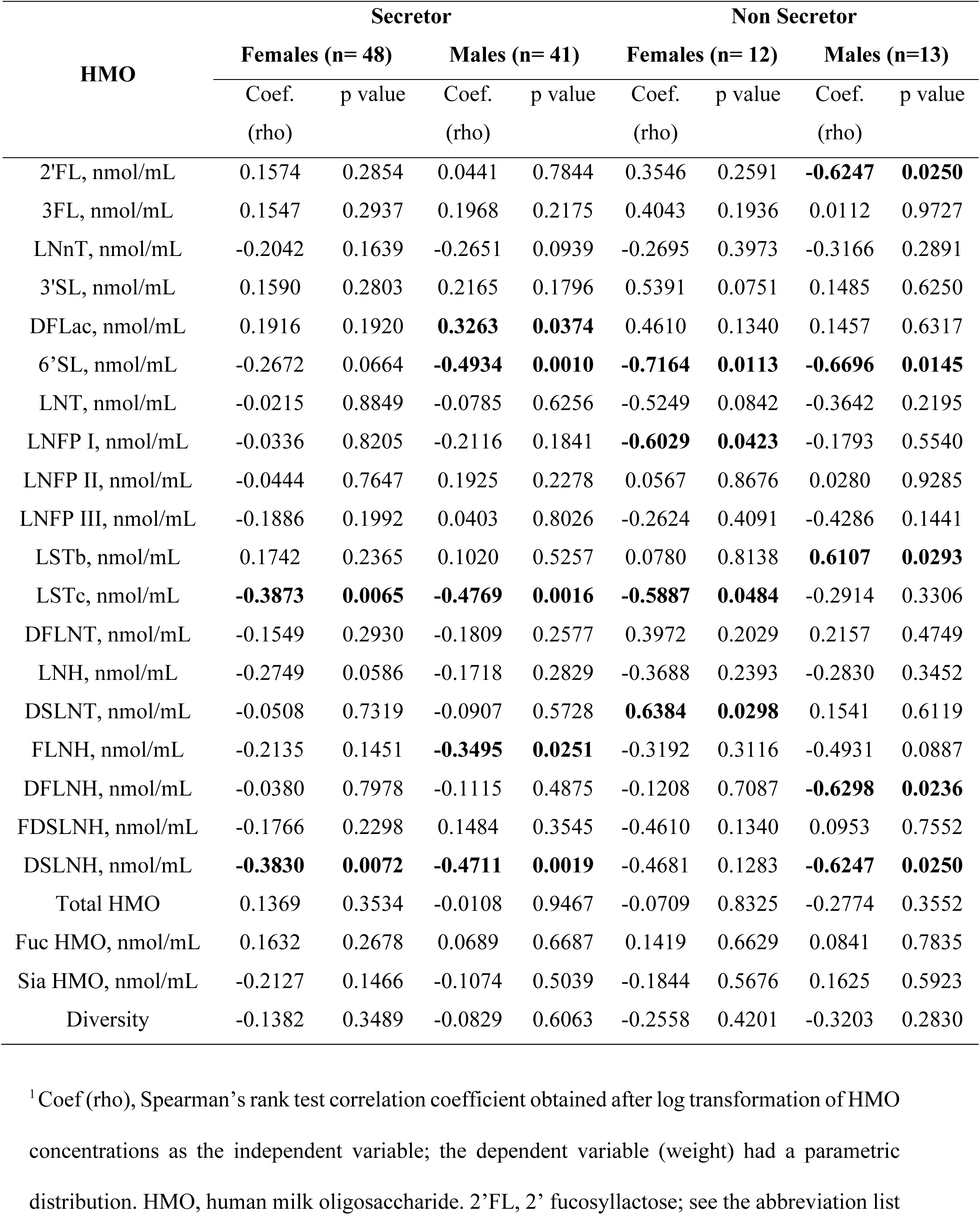
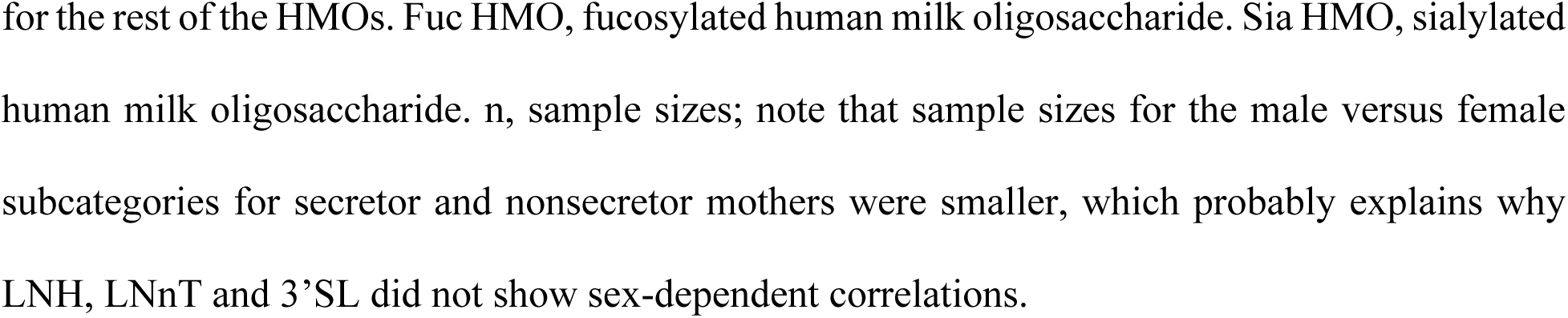
Correlations between HMOs and the weights of female and male children^1^.

| HMO | Secretor |  |  |  | Non Secretor |  |  |  |
| --- | --- | --- | --- | --- | --- | --- | --- | --- |
|  | Females (n= 48) |  | Males (n= 41) |  | Females (n= 12) |  | Males (n=13) |  |
|  | Coef.<br>(rho) | p value | Coef.<br>(rho) | p value | Coef.<br>(rho) | p value | Coef.<br>(rho) | p value |
| 2'FL, nmol/mL | 0.1574 | 0.2854 | 0.0441 | 0.7844 | 0.3546 | 0.2591 | **-0.6247** | **0.0250** |
| 3FL, nmol/mL | 0.1547 | 0.2937 | 0.1968 | 0.2175 | 0.4043 | 0.1936 | 0.0112 | 0.9727 |
| LNnT, nmol/mL | -0.2042 | 0.1639 | -0.2651 | 0.0939 | -0.2695 | 0.3973 | -0.3166 | 0.2891 |
| 3'SL, nmol/mL | 0.1590 | 0.2803 | 0.2165 | 0.1796 | 0.5391 | 0.0751 | 0.1485 | 0.6250 |
| DFLac, nmol/mL | 0.1916 | 0.1920 | **0.3263** | **0.0374** | 0.4610 | 0.1340 | 0.1457 | 0.6317 |
| 6'SL, nmol/mL | -0.2672 | 0.0664 | **-0.4934** | **0.0010** | **-0.7164** | **0.0113** | **-0.6696** | **0.0145** |
| LNT, nmol/mL | -0.0215 | 0.8849 | -0.0785 | 0.6256 | -0.5249 | 0.0842 | -0.3642 | 0.2195 |
| LNFP I, nmol/mL | -0.0336 | 0.8205 | -0.2116 | 0.1841 | **-0.6029** | **0.0423** | -0.1793 | 0.5540 |
| LNFP II, nmol/mL | -0.0444 | 0.7647 | 0.1925 | 0.2278 | 0.0567 | 0.8676 | 0.0280 | 0.9285 |
| LNFP III, nmol/mL | -0.1886 | 0.1992 | 0.0403 | 0.8026 | -0.2624 | 0.4091 | -0.4286 | 0.1441 |
| LSTb, nmol/mL | 0.1742 | 0.2365 | 0.1020 | 0.5257 | 0.0780 | 0.8138 | **0.6107** | **0.0293** |
| LSTc, nmol/mL | **-0.3873** | **0.0065** | **-0.4769** | **0.0016** | **-0.5887** | **0.0484** | -0.2914 | 0.3306 |
| DFLNT, nmol/mL | -0.1549 | 0.2930 | -0.1809 | 0.2577 | 0.3972 | 0.2029 | 0.2157 | 0.4749 |
| LNH, nmol/mL | -0.2749 | 0.0586 | -0.1718 | 0.2829 | -0.3688 | 0.2393 | -0.2830 | 0.3452 |
| DSLNT, nmol/mL | -0.0508 | 0.7319 | -0.0907 | 0.5728 | **0.6384** | **0.0298** | 0.1541 | 0.6119 |
| FLNH, nmol/mL | -0.2135 | 0.1451 | **-0.3495** | **0.0251** | -0.3192 | 0.3116 | -0.4931 | 0.0887 |
| DFLNH, nmol/mL | -0.0380 | 0.7978 | -0.1115 | 0.4875 | -0.1208 | 0.7087 | **-0.6298** | **0.0236** |
| FDSLNH, nmol/mL | -0.1766 | 0.2298 | 0.1484 | 0.3545 | -0.4610 | 0.1340 | 0.0953 | 0.7552 |
| DSLNH, nmol/mL | **-0.3830** | **0.0072** | **-0.4711** | **0.0019** | -0.4681 | 0.1283 | **-0.6247** | **0.0250** |
| Total HMO | 0.1369 | 0.3534 | -0.0108 | 0.9467 | -0.0709 | 0.8325 | -0.2774 | 0.3552 |
| Fuc HMO, nmol/mL | 0.1632 | 0.2678 | 0.0689 | 0.6687 | 0.1419 | 0.6629 | 0.0841 | 0.7835 |
| Sia HMO, nmol/mL | -0.2127 | 0.1466 | -0.1074 | 0.5039 | -0.1844 | 0.5676 | 0.1625 | 0.5923 |
| Diversity | -0.1382 | 0.3489 | -0.0829 | 0.6063 | -0.2558 | 0.4201 | -0.3203 | 0.2830 |

<sup>1</sup> Coef (rho), Spearman's rank test correlation coefficient obtained after log transformation of HMO concentrations as the independent variable; the dependent variable (weight) had a parametric distribution. HMO, human milk oligosaccharide. 2'FL, 2' fucosyllactose; see the abbreviation list
for the rest of the HMOs. Fuc HMO, fucosylated human milk oligosaccharide. Sia HMO, sialylated human milk oligosaccharide. n, sample sizes; note that sample sizes for the male versus female subcategories for secretor and nonsecretor mothers were smaller, which probably explains why LNH, LNnT and 3'SL did not show sex-dependent correlations.

### Associations between HMO concentrations and LAZ/HAZ scores for all children

Children were categorized as short, normal, or tall based on length/height for age-adjusted z (LAZ/HAZ) scores. The relationship between childrens height/length and median concentrations of HMOs in their mothers’ milk were compared by Kruskall-Wallis and Mann‒Whitney U tests. There were significant differences in the median milk concentrations of 7 HMOs and total HMOs fed to short, normal and tall children of all mothers combined, secretor and nonsecretor mothers according to the Kruskal‒Wallis test (**Figure 1 and Supplemental file 1**). According to pairwise comparisons via the Mann‒Whitney U test, compared with tall children, short- and normal-height children of all mothers combined were fed significantly greater concentrations of 3FL and DFLac. In contrast, the tall children of all mothers combined and secretor mothers were fed significantly higher concentrations of 6’SL and LSTc compared to short children of similar mothers; the tall children of secretor mothers were fed significantly greater concentrations of DFLNH and DSLNH than were the short children of similar mothers; and the tall children of nonsecretor mothers were fed significantly greater concentrations of LNnT and total HMOs (**Figure 1 and Supplemental file 1**).

**FIGURE 1.**
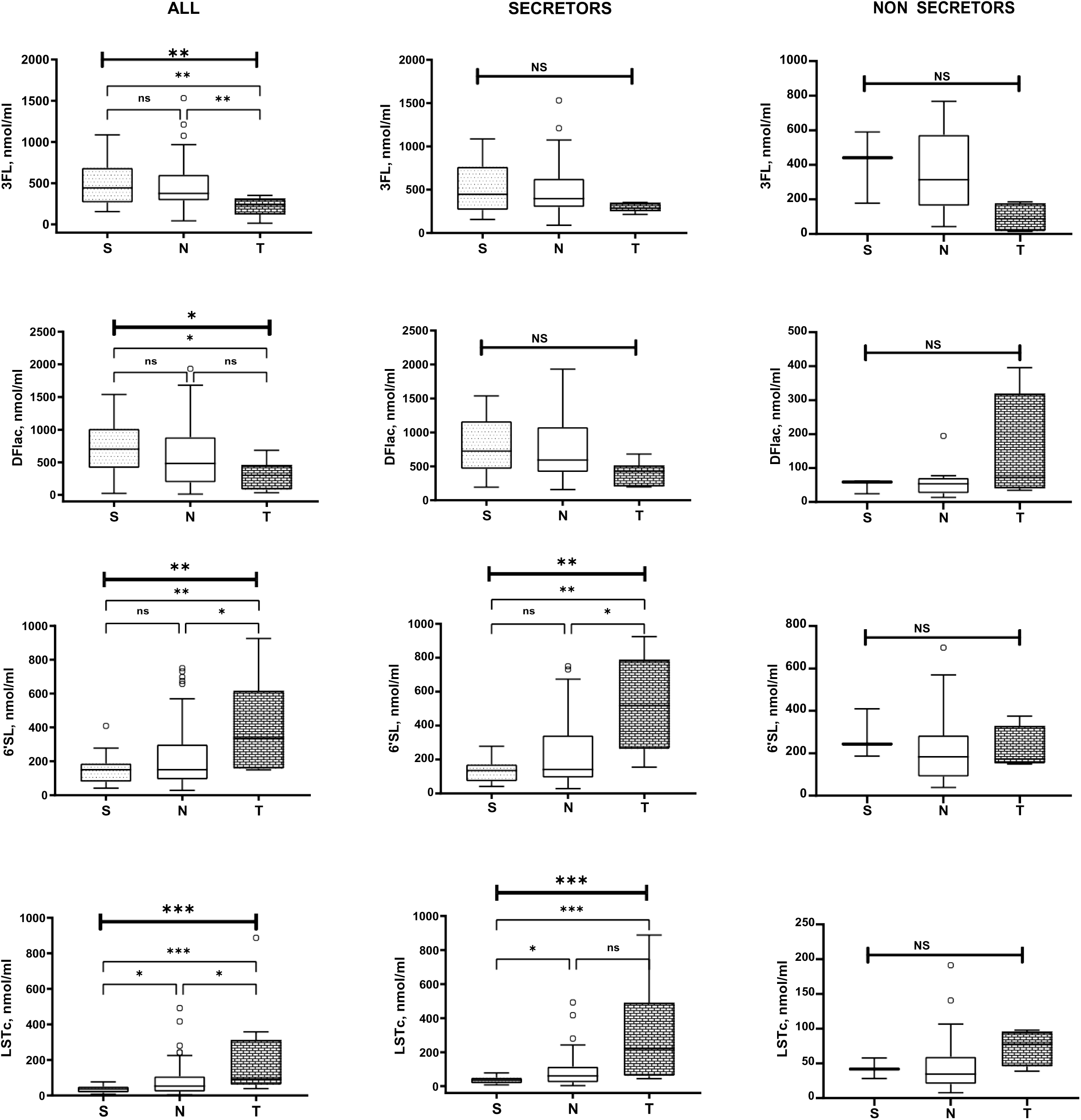
Association between HMO concentrations and LAZ/HAZ scores for all children. S, short children; N, normal-height children; T, tall children. HMO abbreviations are indicated in the list of abbreviations. The data are shown as box and whisker plots with the medians and the lower and upper quartiles or interquartile range (IQR). P values represent the outcomes of comparisons of the three and two LAZ subcategories according to the Kruskal‒Wallis and Mann‒Whitney U tests, respectively. NS, nonsignificant; * P < 0.05; ** P < 0.01; *** P < 0.001.

### Associations between HMO concentrations and WAZ scores in all children

Breastfed children were categorized as underweight, normal or overweight for all mothers combined, and secretor mothers based on weight-for-age-adjusted (WAZ) z scores. There were only two underweight children among the nonsecretor group of mothers. Therefore, comparisons of HMO concentrations were made between all mothers and secretor mothers of underweight, normal and overweight children and between nonsecretor mothers of normal weight and overweight children by the Kruskal‒Wallis test and Mann‒Whitney U test, respectively (**Figure 2, panel A**). The underweight, normal and overweight children of all mothers combined, secretor mothers and nonsecretor mothers were fed significantly different concentrations of 6’SL, LSTc, and LNnT. According to pairwise comparisons, overweight children of all mothers combined, and secretor mothers were fed a significantly higher median concentration of 6’SL and LSTc than were normal weight and underweight children. Lastly, overweight children of secretor mothers were fed a significantly greater median concentration of LNnT compared to underweight children. There were no significant differences in the concentrations of 6’SL, LSTc, or LNnT fed to normal or overweight children of non-secretor mothers, probably because of the small sample sizes of the subcategories.

**FIGURE 2.**
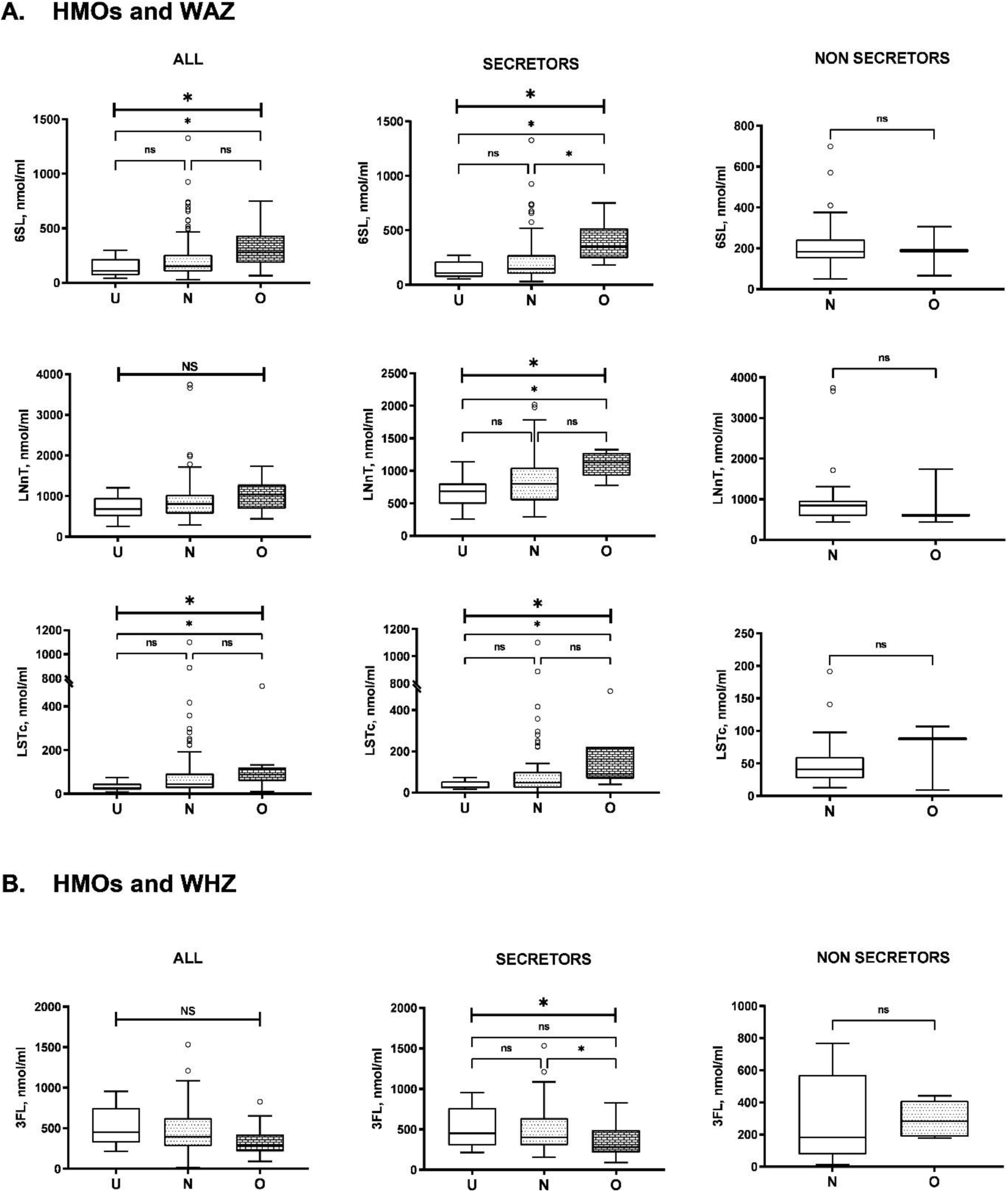
Association between HMO concentrations and WAZ/WHZ scores in all children. U, underweight children; N, normal weight children; O, overweight children. Panel **A**: HMOs and WAZ. Panel **B**: HMOs and WHZ. HMO abbreviations are indicated in the list of abbreviations. The data are shown as box and whisker plots with the medians and the lower and upper quartiles or interquartile range (IQR). P values represent the outcomes of comparisons of the three and two WAZ or WHZ subcategories according to the Kruskal‒Wallis and Mann‒Whitney U tests, respectively. NS, nonsignificant; * P < 0.05.

### Association of HMO concentrations with WHZ scores in all children

Underweight, normal and overweight children of all mothers combined, and secretor but not nonsecretor mothers were fed significantly different concentrations of 3FL (**Figure 2, panel B**). Compared with overweight children, normal weight children of secretor mothers were fed a significantly greater concentration of 3FL. None of the other HMOs had any significant association with the WHZ score.

### Associations between HMO concentrations and LAZ scores in male and female children

A significantly lower 6’SL concentrations were observed in mothers of short- and normal-height female children than in mothers of tall female children (**Figure 3**). No significant differences were detected in mothers of male children. In contrast, significant differences in the LSTc concentration were observed between mothers of both female and male children. According to pairwise comparisons, significantly lower concentrations of LSTc were observed in mothers of short male and female children than in mothers of tall male and female children. The difference in LSTc concentrations between mothers of short- and normal-height children was observed only in mothers of male children. Notably, 5 HMOs and total HMOs, which were significantly associated with LAZ subcategories in all the children considered together, as shown in Figure 1, did not show any associations when the data were reanalyzed by sex.

**FIGURE 3.**
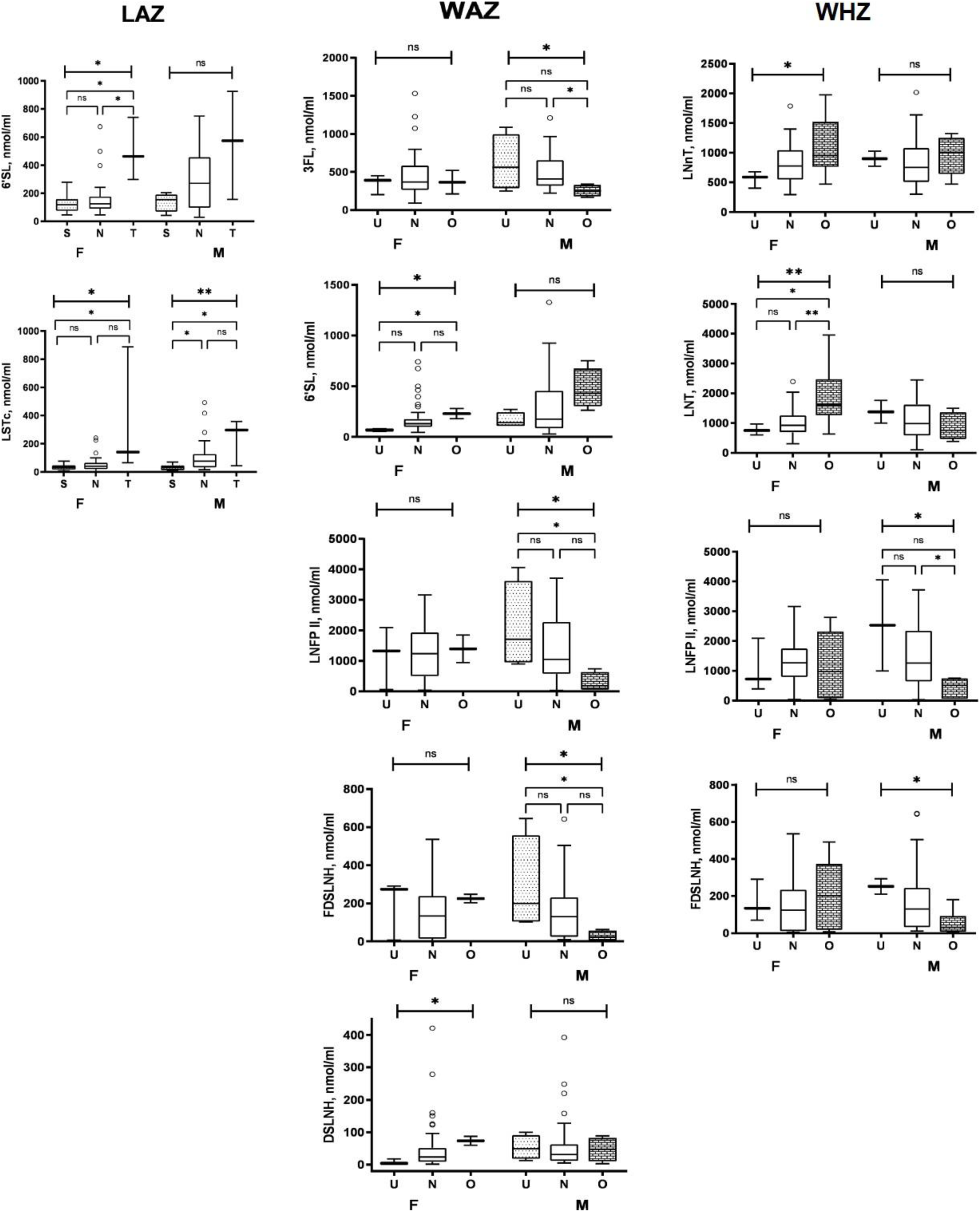
Association between HMO concentrations and LAZ/WAZ/WHZ scores in male and female children. U, underweight children; N, normal weight children; O, overweight children; F, females; M, males. HMO abbreviations are indicated in the list of abbreviations. The data are shown as box and whisker plots with the medians and the lower and upper quartiles or interquartile range (IQR). P values represent the outcomes of comparisons of two LAZ, WAZ or WHZ subcategories according to the Mann‒Whitney U test. NS, nonsignificant; * P < 0.05; ** P < 0.01

### Associations between HMO concentrations and WAZ scores in male and female children

First, significantly higher concentrations of 3FL, LNFP II, and FDSLNH were observed in mothers of underweight male children than in mothers of overweight male children (**Figure 3**). No significant differences in these HMOs were observed between mothers of underweight and overweight female children. Second, significantly higher concentrations of 6’FL and DSLNH were observed in mothers of overweight female children compared to those in mothers of underweight female children. Notably, associations with WAZ subcategories were observed only for LNFPII, FDSLNH and DSLNH when the data were analyzed by sex but not when all the children were considered together.

### Association between HMO concentrations and WHZ scores in male and female children

Significantly higher concentrations of LNnT and LNT were observed in secretor mothers of overweight female children compared to those in secretor mothers of underweight and, for LNT, normal weight female children (**Figure 3**). No significant differences in both HMOs were observed between the secretor mothers of male children. Significantly higher concentrations of LNFP II and FDSLNH were observed in secretor mothers of underweight and overweight male children (**Figure 3**). No significant differences in either HMO were observed between secretor mothers of female children. The associations with WHZ subcategories were observed only for LNnT, LNT, LNFPII, and FDSLNH when the data were analyzed by sex but not when all the children were considered together. Notably, 3FL, which was significantly associated with the WHZ subcategory in mothers of secretor children, did not significantly differ between males and females, probably because of the small sample sizes when the data were subcategorized by sex.

## Discussion

The World Health Organization recommends breastfeeding children for up to 2 years of age for healthy growth and development since human milk contains nutrients, growth factors, and immunological factors required for healthy growth and development [32]. HMOs constitute the third most abundant component of human milk after lactose and lipids, and provide protection against infections and affect growth and cognitive development in breastfed children [1]. Here, we report that in human milk samples collected from breastfeeding Ugandan mothers at a single time point postpartum, representing a wide range of lactation durations spanning 3-103 weeks, differences in the composition and concentrations of HMOs were associated with differences in the height and weight of their breastfed children. We also report for the first time that the association between some HMOs and growth depends on the sex of the children in addition to maternal secretor status.

There were positive or negative Spearman correlations between concentrations of specific HMOs and the heights and weights of children depending on the mothers’ secretor status. Higher concentrations of specific HMOs were fed to children with different height (LAZ) or weight (WAZ and WHZ) categories depending on mothers’ secretor status. Tall stature was associated with higher concentrations of 6’SL, LSTc, DFLNH, DSLNH, LNnT and total HMOs in secretor or nonsecretor mothers; short stature was associated with higher concentrations of 3’FL and DFLac; overweight children of secretor and nonsecretor mothers were fed on higher concentrations of 6’SL, LSTc and LNnT, respectively; and normal weight-for-length (WHZ) was associated with higher concentrations of 3’FL in human milk. Some HMOs had significant correlations with height or weight but showed no significant associations when these growth measurements were transformed into LAZ and WAZ scores; the lack of significance is probably due to the small sample size of mother–child pairs. Finally, the associations between specific HMOs and height and weight or LAZ, WAZ and WFL subcategories significantly differed between male and female children depending on mothers’ secretor status, suggesting sexual dimorphism in HMO-induced growth. Sex differences in the gut microbiota composition have been reported in children under 5 years old [33], but there is a dearth of information about sex-specific metabolism of HMOs by the gut microbiota. To the best of our knowledge, no previous studies have reported sex differences in the association between HMOs and growth.

Similar studies in the USA and Europe demonstrated an association between HMO concentrations or uptake and growth in children, with 2’FL being positively associated with and LNnT and LNFPI being negatively associated with children’s height and weight [20,21,22]. These studies did not report sex differences. In our Ugandan mother–child pairs, 2’FL and LNFP I-secretor-dependent HMOs did not have a significant effect on weight in children of secretor mothers but were negatively correlated with weight in children of nonsecretor mothers. A Gambian study that analyzed the HMO composition and infant gut microbiota of mother–infant pairs at 4, 16 and 20 weeks after delivery demonstrated that LNFPI was predictive of the HAZ score at 20 weeks, while 3’SL was predictive of infant WAZ score [23]. In a Malawian study, milk samples were collected at a single time point at 6 months postpartum, after which the growth and development endpoints were longitudinally monitored for more than one year [24]. There was a positive association between the total abundance of HMOs and the change in LAZ from 6 to 12 months and between total HMOs, HMO-bound fucose and sialic acid and language development at 18 months among secretors. Whereas in the Gambian study, LNFPI + LNFPIII was predictive of HAZ, in our study, 6’SL, LSTc and three other HMOs and total HMOs were significantly associated with increased HAZ (tall stature). Again, unlike in the Gambian study, 3’SL was not associated with growth in Ugandan infants and children, but 3’FL was negatively associated with WHZ. Our study identified 6’SL and LSTc as important determinants of HAZ, which were not reported in Gambian children. In the Malawian study, 6’SL and 3’SL were negatively and positively associated with changes in the head circumference Z score. Our study did not measure head circumference. The Gambian study highlighted the effect of season with higher HMO production during the dry season. Our study measured HMOs in human milk collected at a single time point at the beginning of the rainy season, but the mothers were not longitudinally followed up. The differences in outcomes between our study and those of the Gambian, Malawian, and Western studies might be due to differences in study designs and populations, diets, seasons of study, and geography.

Although numerous studies have reported significant associations between specific HMOs and the height and weight of breastfeeding children [20,21,22,23,24], none of the studies have reported differences in these associations between male and female children. In this study, stratifying the data by sex revealed significant associations between height and weight and specific HMOs in male and female children, which would otherwise have remained obscured. HMO-bound fucose and seven HMOs, 2’FL, LNFP I, 3FL, DFLNH, DSLNT, LNT and LSTb, exhibited significant associations in male or female children of secretor or nonsecretor mothers only when the data were disaggregated by sex. Our data suggest that maternal secretor status and the sex of breastfeeding children are important factors that must be considered when assessing the association between HMOs and growth measurements.

The mechanisms by which HMOs affect infant stature and weight in general and in a sex-specific manner in particular are not clear given the observational nature of our study. We hypothesize that differences in the microbiota composition between male and female children of secretor and nonsecretor mothers may lead to the observed sex differences in the association between HMOs and growth parameters. First, it has been speculated that HMOs affect growth through prevention of morbidity [24] since HMOs promote specific microbiota whose metabolites directly modulate immune functions [28,34]. It is plausible that, in our study population, HMOs affected children’s growth through microbiota-induced modulation of immunity, which reduced morbidity and thereby promoted changes in HAZ, WAZ and WHZ scores, as proposed [35]. Second, the associations between maternal secretor status and infant microbiota composition [35,36] and between infant microbiota and infant growth [25] may explain the observed differences in the associations between some HMOs and weight and height in children of secretor and nonsecretor mothers. Finally, there are sex differences in the gut microbiota composition of male and female children under 5 years old [33,38,39,40,41], which probably determines how the microbiota and HMOs interact to regulate growth in male and female Uganda children. These hypotheses should be tested in hypothesis-driven mechanistic longitudinal studies of mother–child pairs and randomized controlled clinical trials of commercially available HMOs used as food supplements. The outcomes of these studies may inform HMO combinations that can be used as food supplements for African children who may not enjoy the nutritional benefits of breastfeeding and the design of clinical trials assessing the growth and immunological effects of HMO supplementation.

This study has several limitations. First, the small sample size and the observational nature of the study did not allow us to perform statistical modeling to adjust for potential confounders (e.g., complementary feeding, maternal age and BMI, mode of delivery) or to perform multiple correction tests. The mode of delivery was probably not an important confounder since there were practically no Caesarian sections among the Ugandan mothers. Second, the collection of human milk at only one time point could have missed important changes in HMO composition during lactation [42,43]. However, we believe that our study cross-sectionally captured HMO compositional changes during lactation because the mothers represented a range of lactation stages from 3 to 103 weeks, which is greater than what has been reported in other similar studies. Third, anthropometric measurements in children were carried out at a single time point during sample collection, and the lack of serial measurements made it impossible to assess causal relationships between growth velocity and various HMOs. Fourth, the study did not collect fecal samples for analysis of microbiota composition and therefore cannot mechanistically link the observed children’s growth outcomes to the effect of HMOs on microbiota composition. Finally, information about feeding practices was not collected for the Ugandan children in this study, yet diet may affect the interaction between sex and the microbiota [33,41].

This study has several strengths despite its cross-sectional nature. First, for the first time, we demonstrated sexual dimorphism in the relationship between specific HMOs and growth parameters in children. These sex differences were discovered via post hoc analyses and remain to be investigated in future mechanistic studies. These studies could inform future nutritional interventions involving combinations of HMOs as food supplements that are equally effective for both male and female children. Second, this study examined the association between HMOs and growth in a wider range of children’s ages and lactation durations than did previous studies. Finally, the weights and heights of the children and mothers were not self-reported but were measured by trained research staff using calibrated standard equipment.

### Perspective and Significance

Our observational study demonstrated for the first time sex differences in the association between high concentrations of some HMOs and stature and weight in Ugandan children of secretor and nonsecretor mothers. Our study highlights the usefulness of analyzing these associations using data grouped by sex since considering sex-disaggregated data revealed an association with several HMOs, which was not evident otherwise. The sex differences were not detected during post hoc analyses and must be validated through hypothesis-driven mechanistic longitudinal studies of HMO concentrations and intake and growth in mother–child pairs, as well as randomized controlled trials of commercially available HMOs as food supplements in children. These validation studies should factor the interactions between maternal secretor status and the sex of breastfeeding children as important covariates. We believe that if validated, these results may be generalizable to breastfeeding infants and children in other countries and may inform sex-specific nutritional interventions that employ combinations of HMOs as infant food supplements.

## Data Availability

The datasets supporting the conclusion of this article will be made available upon request from TGE.

## Abbreviations

BMI: body mass index
FUT2: α1,2-fucosyltransferase gene
HMO: human milk oligosaccharide
LAZ: height/length for age Z score
MUAC: mid-upper arm circumference
WAZ: weight for age Z score
WHZ: weight-for-height/length Z score

## List of HMO abbreviations

2’FL: 2’fucosyllactose
3 FL: 3-fucosyllatose
3’SL: 3’sialyllactose
6’SL: 6’sialyllactose
DFlLac: difucosyllactose
DFLNH: Difucosyllacto-N-hexaose
DFLNT: Difucosyllacto-N-tetrose
DSLNH: disialyllacto-N-tetraose.
DSLNT: Disialyllacto-N-tetraose
FDSLNH: Fucosyldisialyllacto-N-hexaose
FLNH: Fucosyllacto-N-hexaose
LNFPI: lacto-N-fucopentaose I
LNFPII: lacto-N-fucopentaose II
LNFPIII: lacto-N-fucopentaose III
LNH: Lacto*-*N*-*hexaose
LNnT: Lacto-N-neotetraose
LNT: Lacto-N-tetraose

LSTb Sialyllacto-*N*-tetraose b

LSTc sialyllacto-N-tetraose c

## Declarations

### Ethical consideration

The Uganda National Council for Science and Technology and the Research Ethics Committee of the Vector Division, Ministry of Health, approved the original HD4MC study. All mothers who donated milk samples and brought infants to the study clinic for weight and height measurements and health assessment signed or thumb-printed an informed consent form.

### Consent for publication

Not applicable.

### Competing interest

The authors have no competing interest to disclose.

### Funding

Thomas Egwang received funding from the Global Innovation Fund, and Grand Challenges Canada. The funders played no role in the study design, data collection and analysis, preparation of the manuscript or decision to publish this paper.

### Authors’ contributions

TJO and EO conducted the study, including sample collection; TJO drafted the manuscript; SM and CY performed the HMO laboratory analysis; VIM performed the statistical analysis; TGE and LB conceived and directed the study. All the authors read and approved the final manuscript.

## Acknowledgments

We are very grateful to the Ugandan mother–child pairs for their participation. We would like to thank the clinical and laboratory staff of Med Biotech Laboratories at St Anne HC III Usuk, Katakwi, Uganda.

## Authors Information

LB is University of California San Diego Chair of Collaborative Human Milk Research endowed by the Family Larsson-Rosenquist Foundation (FLRF), Switzerland.

## Supplemental File 1 (pdf)

**Title:** Concentrations of HMOs in breast milk and LAZ subcategories in infants.

**Description of the data:** S, short ; N, normal height children; T, tall children. The data are shown as box and whisker plots with the medians and the lower and upper quartiles or interquartile range (IQR). P values represent the outcomes of comparisons of the three and two LAZ subcategories according to the Kruskal‒Wallis and Mann‒Whitney U tests, respectively. NS, nonsignificant; * P < 0.05.

## Notes

### Competing Interest Statement

The authors have declared no competing interest.

