## Supplementary figures and images for "Sex differences in the associations of human milk oligosaccharides with height and weight in breastfed Ugandan children"

### Supplemental File 1

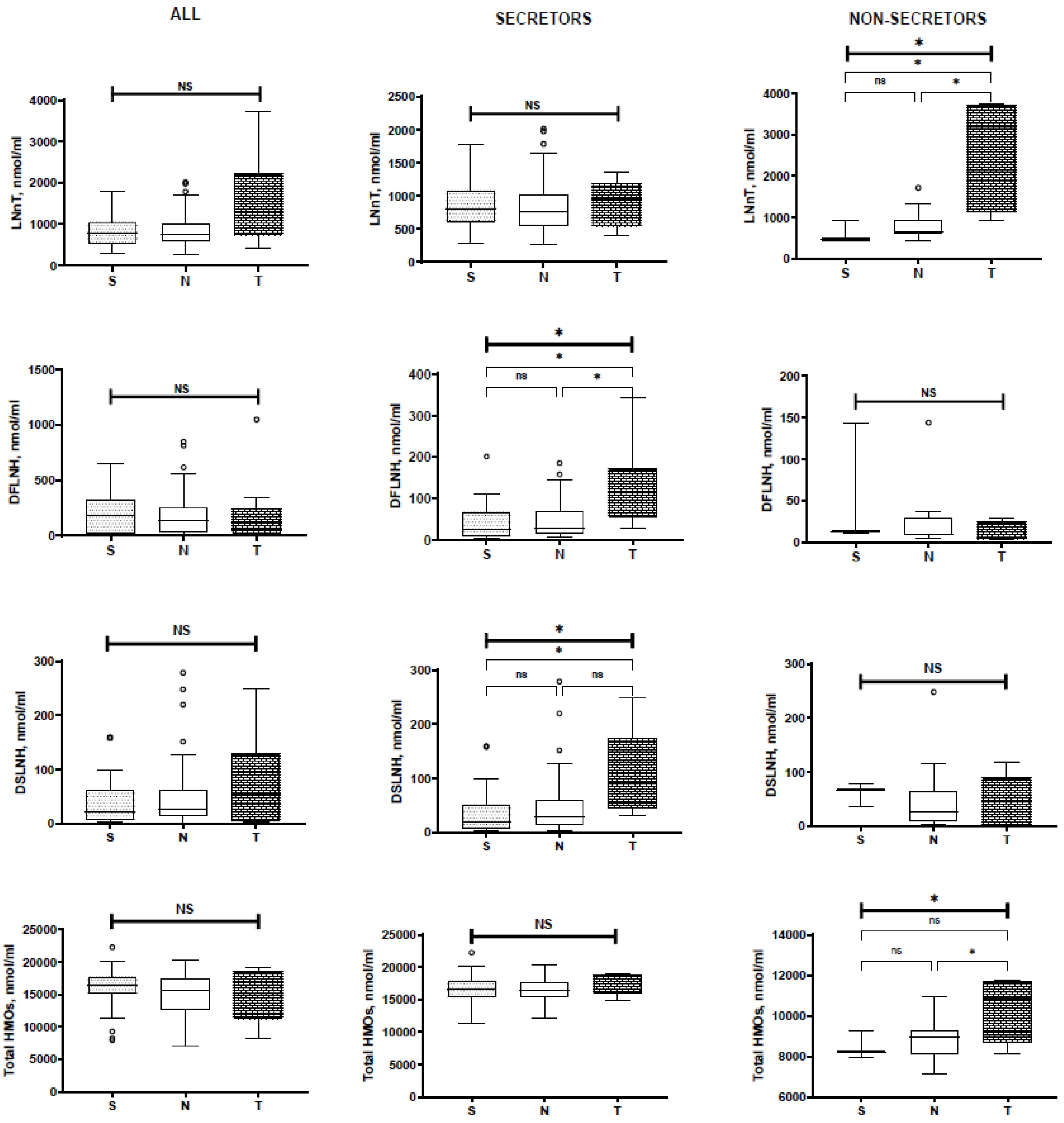
